# Double trouble? When a pandemic and seasonal virus collide

**DOI:** 10.1101/2020.03.30.20047993

**Authors:** Casey Zipfel, Vittoria Colizza, Shweta Bansal

**Affiliations:** Department of Biology, Georgetown University, Washington DC, USA; INSERM, Sorbonne Université, Pierre Louis Institute of Epidemiology and Public Health, Paris, France

**Author notes:** Corresponding author: Casey Zipfel, Shweta Bansal.

**Keywords:** COVID-19, influenza, nonpharmaceutical intervention, vaccination, United States, Europe

## Abstract

As healthcare capacities in the US and Europe reach their limits due to a surge in the COVID-19 pandemic, both regions enter the 2020-2021 influenza season. Southern hemisphere countries that had suppressed influenza seasons provide a hopeful example, but the lack of reduction in influenza in the 2019-2020 influenza season and heterogeneity in nonpharmaceutical and pharmaceutical interventions show that we cannot assume the same effect will occur globally. The US and Europe must promote the implementation and continuation of these measures in order to prevent additional burden to healthcare systems due to influenza.

## Introduction

The COVID-19 pandemic has altered almost every aspect of public health since its global spread in early 2020. The impacts of COVID-19 continue to unfold and one such consequence is the effect of the pandemic on seasonal respiratory pathogens like influenza. The similarities between the transmission pathways and clinical manifestations of SARS-CoV-2 and influenza lead to critical ecological, public health and clinical interactions (Solomon et al., 2020). As the influenza season begins in temperate, northern hemisphere regions, there is a devastating surge of COVID-19 in the US and Europe that is pushing healthcare capacity to its limits, leaving no room for additional public health burden. On the other hand, reports of a curtailed 2019-2020 flu season in the US and a suppressed 2020 flu season in southern hemisphere nations have provided encouraging signs. With the competing forces of viral interactions, overlapping protective behaviors and a single, limited healthcare capacity, it remains unclear what the upcoming winter will bring for influenza.

To understand the impacts of COVID-19 on influenza, we first look retrospectively. By the time SARS-CoV-2 transmission had become widespread in early 2020, the influenza season in Europe was largely over (Boelle, 2020), but flu transmission in the US was still ongoing and declined rapidly in March and April (CDC, 2020a). Some have proposed that the pandemic and the ensuing nonpharmaceutical interventions led to this unexpected decrease in influenza transmission (Olsen et al., 2020). However, the viral circulation in the 2019-2020 season and the timing of nonpharmaceutical interventions make these impacts unclear.

Global case studies provide further evidence to assess the impacts of COVID-19 on influenza transmission. The countries in the southern hemisphere, which typically experience their influenza season in May-October, have reported drastically reduced influenza circulation in 2020 (Olsen et al., 2020). One possible explanation for this pattern is viral competition between SARS-CoV-2 and influenza. This competition could occur through multiple mechanisms, such as immune interactions, viral competition and a reduced susceptible pool due to isolation (Nickbakhsh et al., 2019; Rohani et al., 2003). But the more likely explanation is the significant behavioral and pharmaceutical interventions in place in these settings that are leading to this dramatic effect on flu. Nonpharmaceutical interventions such as closures of non-essential businesses, telework, restriction on gathering size, and mask-wearing have been key public health tools for limiting the impact of the COVID-19 pandemic (Di Domenico et al., 2020; Haug et al., 2020). Given the shared transmission route between SARS-CoV-2 and influenza, the same protective behaviors could greatly limit influenza transmission (Cowling et al., 2020). In fact, far less mitigation effort is required to control a low R_0_ disease like flu (R_0_ = 1-2), compared with a high R_0_ disease like COVID-19 (R_0_ = 2-5). Additionally, increased influenza vaccination is another tool that could reduce influenza burden. Global case studies examining this data can shine light on how these factors have impacted or will impact influenza transmission.

To further our retrospective and prospective understanding of influenza dynamics during the COVID-19 pandemic, we consider case studies across geographic regions. We analyze influenza data from the 2019-2020 season to identify whether COVID-19 significantly impacted influenza transmission in the US. Then, we present global data on mobility and vaccination, for countries that have already avoided an influenza season in the Southern hemisphere and countries that are in the midst of the influenza season in the Northern hemisphere.

## Methods

### Intervention Analysis

We tested whether the 2019-2020 influenza A season was statistically different from expectations based on influenza seasons of prior years. We separate influenza A from influenza B as the influenza B dynamics in 2019-2020 were extremely unusual, mostly occurring early in the season before COVID-19 was a factor in the US, and this obscures comparison of influenza data with prior seasons (Figure S1). We obtained surveillance data from the CDC for the 2015-2016 through the 2019-2020 influenza seasons (Centers for Disease Control and Prevention, 2020). We calculated the influenza A ILI rate as % positive influenza A * ILI / Total visits. This accounts for influenza-like illness cases, healthcare seeking, and influenza A virus circulation. We used intervention analysis as implemented in the CausalImpact R package to build an expectation for the 2019-2020 influenza season, based on the 2015-2016, 2016-2017, 2017-2018, and 2018-2019 seasons as control data (Brodersen et al., 2015). The date of the intervention was March 16, 2020, which is the same week as the COVID-19 National Emergency Declaration in the US. This means that the pre-intervention period was 10/7/2019-3/16/2020, and the post-intervention period was 3/23/2020-7/1/2020. We compare the expected 2019-2020 influenza A season to the actual 2019-2020 influenza A season. We also gathered mobility data from Safegraph to evaluate mobility in this timeframe in the US, shown in Figure S2 (Safegraph, 2020).

### Global intervention comparison

We gathered global mobility data from Google mobility reports (Google, 2020). This represents the effectiveness of non-pharmaceutical interventions for COVID-19 in reducing mobility. We particularly evaluate mobility to retail and recreational locations compared to a pre-pandemic baseline. We also gathered 2020 influenza vaccination data for uptake of influenza vaccination. This was gathered from Reseau Sentinelles and IQVIA France, the CDC, and the Australian Department of Health (CDC, 2020b; Hunt, 2020). As a case study, we compared Australia, France, and the US, as they are westernized, large population countries with data available on these two factors. Australia provides a retrospective example, as it experienced a suppressed influenza season in May-October 2020. We also examined NPIs in other southern hemisphere countries for support (Figure S3). France and the US provide prospective examples for comparison.

## Results

### The 2019-2020 influenza season in the US was not significantly impacted by COVID-19

When examining influenza A dynamics and controlling for healthcare seeking, the dynamics of the 2019-2020 influenza season were not significantly different from what was expected after March 16 based on the 4 prior influenza seasons. It is necessary to isolate influenza A from influenza B because influenza B cases occurred much earlier and at a higher rate than previous years (Figure S1). The high, early influenza B peak makes the 2019-2020 season appear to be very severe early in the season. It also makes comparing the tail of the season misleading: in typical years influenza B becomes more prevalent at the end of the season resulting in a more gradual tail, and its early decrease in 2019-2020 makes the tail of the 2019-2020 season appear to drop more drastically than typical years. Examining just influenza A dynamics reveals similar dynamics to previous years. This is supported by evaluating mobility data in the US in early 2020 (Figure S2). This demonstrates that mobility did not deviate from previous years until April, when the influenza season had ended.

### Southern Hemisphere countries engaged in reduced mobility and increased vaccination during the influenza season

Mobility to retail and recreational locations in Australia was at 30-45% below baseline at the start of the influenza season (Figure 1B). Similar reduced mobility patterns at the start of the influenza season were observed in Chile, Argentina, and South Africa (Figure S3). All of these countries have reported diminished influenza seasons (Olsen et al., 2020; PAHO PHE/IHM/Influenza Team, 2020). Additionally, Australia reported a 36% increase in influenza vaccine distribution in 2020.

**Figure 1.**
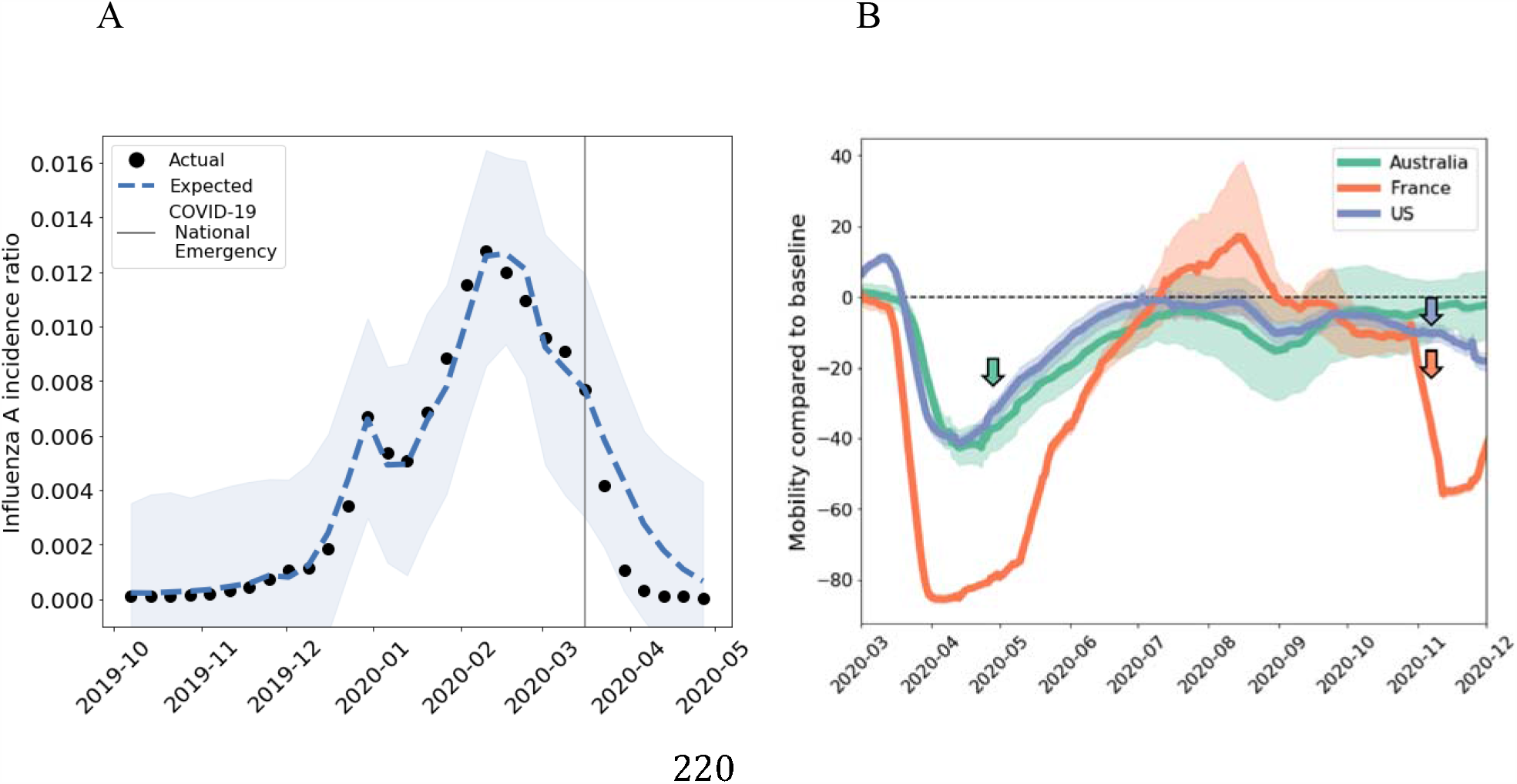
(A) The 2019-2020 influenza season in the US showed no significant impact of COVID-19 pandemic mitigation measures. The weekly influenza A incidence ratio (% positive for influenza A^*^ ILI cases/Total visits, black dots, data from CDC 2020a) compared to what is expected based on the 4 prior influenza seasons (blue dashed line with shaded 95% confidence interval). After the COVID-19 National Emergency declaration in the US (gray line), the influenza epidemic did not deviate significantly from previous seasons. (B) Mobility to retail and recreational locations compared to baseline in Australia (green), France (orange), and the US (green), from Google Community Mobility reports (Google 2020). The lines represent the regional mean for each country, and the shaded region is the standard error. Arrows indicate the typical start of the influenza season for each region in the corresponding color.

### Participation of Northern Hemiphere countries in nonpharmaceutical interventions remains variable during the early influenza season

Mobility to retail and recreational locations at the start of the 2020-2021 influenza season was at 35-40% below baseline. Additionally, vaccine purchase data shows that influenza vaccine distribution was 13% higher in 2020 than in previous years. Mobility to retail and recreational locations in the US at the start of the 2020-2021 influenza season was only slightly below baseline values. Additionally, the US reports an 11% increase in influenza vaccine distribution. However, stark heterogeneities exist both spatially and demographically. The US state of Iowa estimates a decline in vaccination across populations, with a 24 percentage points decline in vaccination in the 65+ age group (Iowa Department of Public Health, 2020). Additionally, it is estimated that vaccination coverage among Black, non-Hispanic children in the US is down 11 percentage points from last year (CDC, 2020b).

## Discussion

Our study does not seek to identify causal or correlative links between influenza and COVID-19 dynamics. Instead, we seek to synthesize various lines of evidence that allow for careful consideration of this relationship in the future. We have considered case studies to explore the impact of the COVID-19 pandemic on influenza dynamics in northern and southern hemisphere nations. We find that the 2019-2020 influenza season in the US was not significantly impacted by COVID-19. We compare mobility and flu vaccine distribution data for Australia, France, and the US. Australia provides an example of suppressed influenza epidemics, in which significantly reduced decreased mobility and increased vaccine distribution was observed during the early part of the influenza season. France appears to be similar to Australia, with reduced mobility and an increase in influenza vaccination. The US, however, has mobility similar to pre-pandemic baseline, and an increase in vaccination, with notable spatial and demographic heterogeneity.

Influenza surveillance in the US through November 30th shows lower rates of disease compared to previous seasons (Centers for Disease Control and Prevention, 2020), and Europe has reported only 5 sentinel-source positive specimens out of 8729 samples through December 6^th^ (Figure S4, ECDC-WHO, 2020). Travel restrictions due to the pandemic may also limit the introduction of influenza from other countries. This encouraging news must be balanced with caution. Healthcare systems are stretched to capacity with current COVID-19 hospitalizations topping 113,000 in the US and 140,000 in Europe, so we cannot afford additional burden from influenza (which leads to hundreds of thousands of hospitalizations annually in the US and Europe). Recent data also suggests that while the risk of testing positive for SARS-CoV-2 was 68% lower among influenza cases, the risk of severe outcomes, such as ICU admission or death, were many-fold higher for coinfection cases than for either infection alone (Stowe et al., 2020). Thus, though current flu numbers are fortunate, continued diligence and public health messaging will be necessary to continue to suppress influenza cases.

The lessons of the COVID-19 pandemic must be heeded to prevent further devastation. We must sustain efforts towards social distancing and mask wearing, which have been definitively linked to reductions in respiratory disease transmission. We must continue vaccination administration efforts while accommodating social distancing requirements to prevent exacerbation of health disparities. We must not let hopeful news of suppressed epidemics deter from a path of action and vigilance.

## Data Availability

All data used in the study is publicly available and cited in the work

https://gis.cdc.gov/grasp/fluview/fluportaldashboard.html

https://www.google.com/covid19/mobility/

## Ethics Statement

This study and use of the SafeGraph dataset was approved by the Georgetown-Medstar IRB with study ID STUDY00003041.

## Acknowledgments

We thank Reseau Sentinelles and IQVIA France for providing us access to flu vaccine sale data. Research reported in this publication was supported by the National Institute Of General Medical Sciences of the National Institutes of Health under Award Number R01GM123007. The content is solely the responsibility of the authors and does not necessarily represent the official views of the National Institutes of Health. We also acknowledge support from the PhRMA Foundation.

## References

Boelle P et al. Excess cases of Influenza like illnesses in France synchronous with COVID19 invasion 2020.

Brodersen KH, Gallusser F, Koehler J, Remy N, Scott SL. Inferring causal impact using bayesian structural time-series models. Ann Appl Stat 2015;9:247–74. https://doi.org/10.1214/14-AOAS788.

CDC. FluView Summary ending on September 26, 2020. FluView 2020a. https://www.cdc.gov/flu/weekly/weeklyarchives2019-2020/Week39.htm.

CDC. Weekly National Flu Vaccination Dashboard. Wkly Natl Flu Vaccin Dashboard 2020b. https://www.cdc.gov/flu/fluvaxview/dashboard/vaccination-dashboard.html.

Centers for Disease Control and Prevention. Weekly U.S. Influenza Surveillance Report 2020. https://www.cdc.gov/flu/weekly/index.htm.

Cowling BJ, Ali ST, Ng TWY, Tsang TK, Julian CM. Impact assessment of non-pharmaceutical interventions against COVID-19 and influenza in Hong Kong: an observational study 2020.

Di Domenico L, Pullano G, Sabbatini CE, Boëlle PY, Colizza V. Impact of lockdown on COVID-19 epidemic in Île-de-France and possible exit strategies. BMC Med 2020;18:1–13. https://doi.org/10.1186/s12916-020-01698-4.

ECDC-WHO. Primary Care Data. Flu News Eur 2020. https://flunewseurope.org/PrimaryCareData/SentinelVirologicalDetections.

Google. Community Mobility Reports 2020. https://www.google.com/covid19/mobility/.

Haug N, Geyrhofer L, Londei A, Dervic E, Desvars-Larrive A, Loreto V, et al. Ranking the effectiveness of worldwide COVID-19 government interventions. Nat Hum Behav 2020;4. https://doi.org/10.1038/s41562-020-01009-0.

Hunt G. Record flu vaccines in 2020 to protect Australians. Dep Heal 2020. https://www.health.gov.au/ministers/the-hon-greg-hunt-mp/media/record-flu-vaccines-in-2020-to-protect-australians.

Iowa Department of Public Health. Influenza Vaccine Data. Iowa Public Heal Track Portal 2020. https://tracking.idph.iowa.gov/Health/Immunization/Influenza-Vaccine/Influenza-Vaccine-Data.

Nickbakhsh S, Mair C, Matthews L, Reeve R, Johnson PCD, Thorburn F, et al. Virus-virus interactions impact the population dynamics of influenza and the common cold. Proc Natl Acad Sci U S A 2019;116:27142–50. https://doi.org/10.1073/pnas.1911083116

Olsen SJ, Azziz-Baumgartner E, Budd AP, Brammer L, Sullivan S, Pineda RF, et al. Decreased Influenza Activity During the COVID-19 Pandemic — United States, Australia, Chile, and South Africa, 2020. MMWR Morb Mortal Wkly Rep 2020;69:1305–9. https://doi.org/10.15585/mmwr.mm6937a6.

PAHO PHE/IHM/Influenza Team. Weekly Influenza Report EW 49. 2020.

Rohani P, Green CJ, Mantilla-Beniers NB, Grenfell BT. Ecological interference between fatal diseases. Nature 2003;422:885–8. https://doi.org/10.1038/nature01542.

Safegraph. Social Distancing Metrics dataset 2020.

Solomon DA, Sherman AC, Kanjilal S. Influenza in the COVID-19 Era. JAMA 2020;324:1342–3. https://doi.org/10.7189/JOGH.10.011101.

Stowe J, Tessier E, Zhao H, Guy R, Muller-Pebody B, Zambon M, et al. Title: Interactions between SARS-CoV-2 and Influenza and the impact of coinfection on disease severity: A test negative design. MedRxiv 2020:2020.09.18.20189647.

Zipfel C, Bansal S. Assessing the interactions between COVID-19 and influenza in the United States. MedRxiv Prepr Serv Heal Sci 2020:1–13. https://doi.org/10.1101/2020.03.30.20047993.

