## Supplement for "Double trouble? When a pandemic and seasonal virus collide"

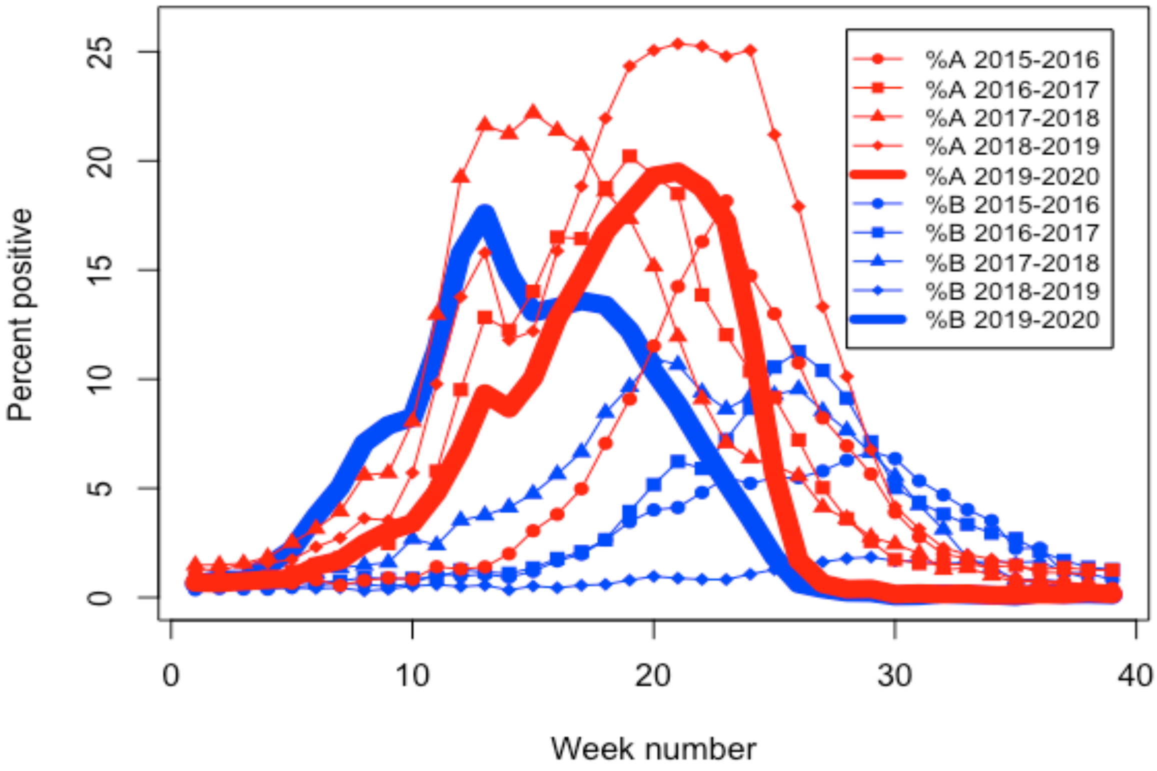


**Figure S1**. Influenza viral circulation in the 2015-2016 season (circle), the 2016-2017 season (square), the 2017-2018 season (triangle), the 2018-2019 season (diamond), and the 2019-2020 season (thick line) from CDC influenza surveillance data (CDC 2020e). The percent positive rate shows that in prior years, the percent positive for influenza A (red) is higher and earlier than the percent positive for influenza B (blue). In the 2019-2020 influenza season, influenza B (thick blue line) occurs much earlier and at a higher rate than usual. The 2019-2020 influenza A (thick red line) looks somewhat similar to prior seasons.

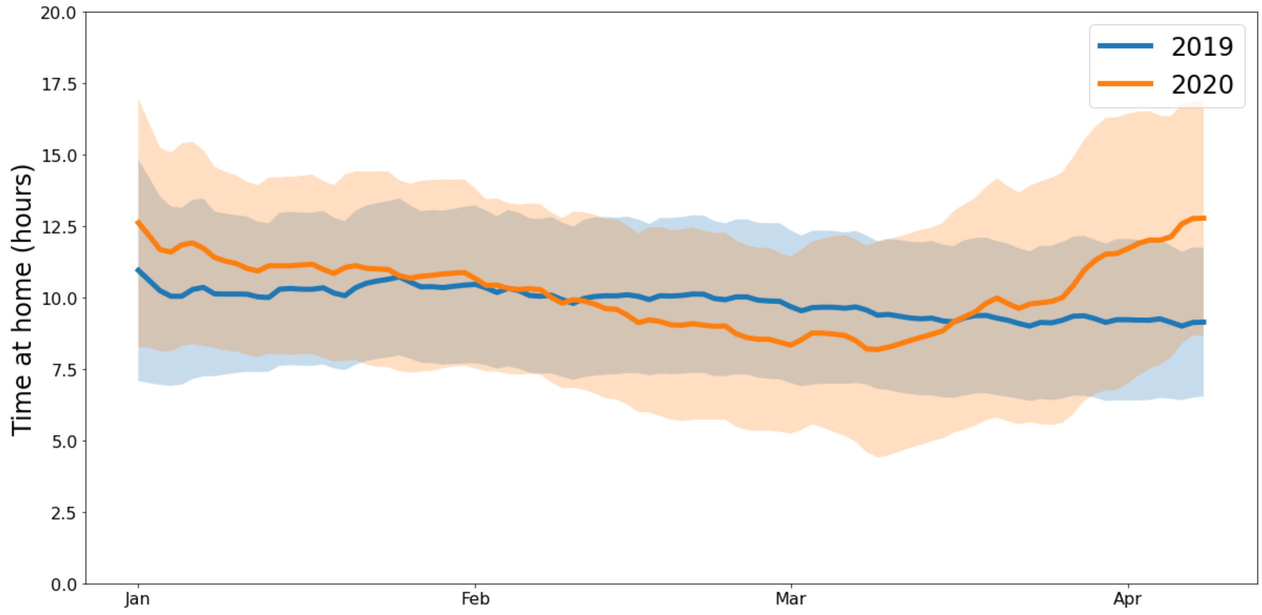


**Figure S2**. Mean and standard deviation of time spent at home in hours in the US in 2019 (blue), and in 2020 (orange). Data from Safegraph[1]. During the timeframe of the 2019-2020 influenza season (January through March), mobility was comparable to prior years.

[1] Safegraph Social Distancing Metrics dataset: https://www.safegraph.com/

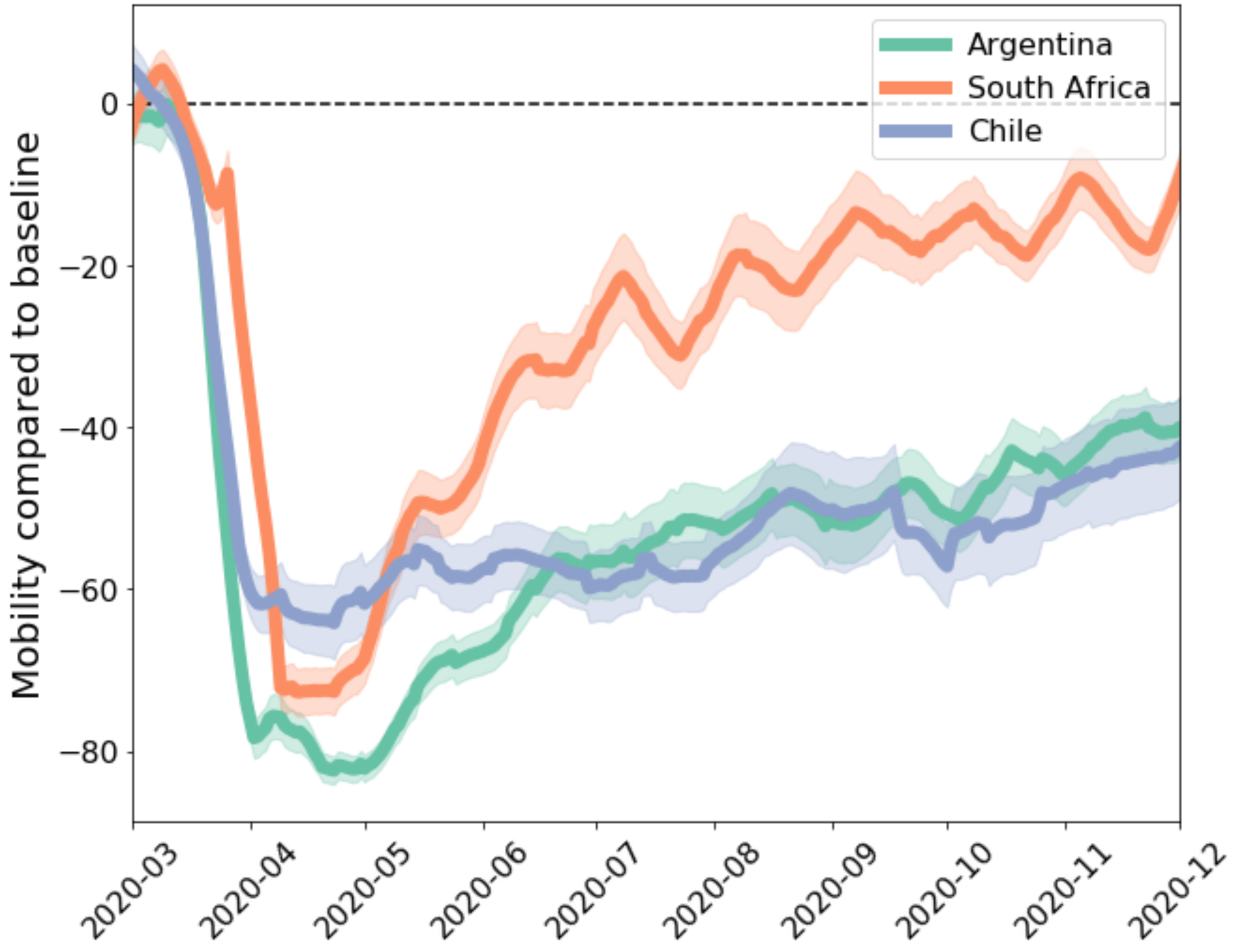


**Figure S3.** Google mobility data for southern hemisphere countries that experienced suppressed influenza seasons in 2020 (Argentia: green, South Africa: orange, and Chile: purple). This is the daily subregion mean and standard error of the mobility to retail and recreational locations compared to baseline. The influenza season in these countries typically occurs in May-September, during which all of the countries have reduced mobility compared to baseline.

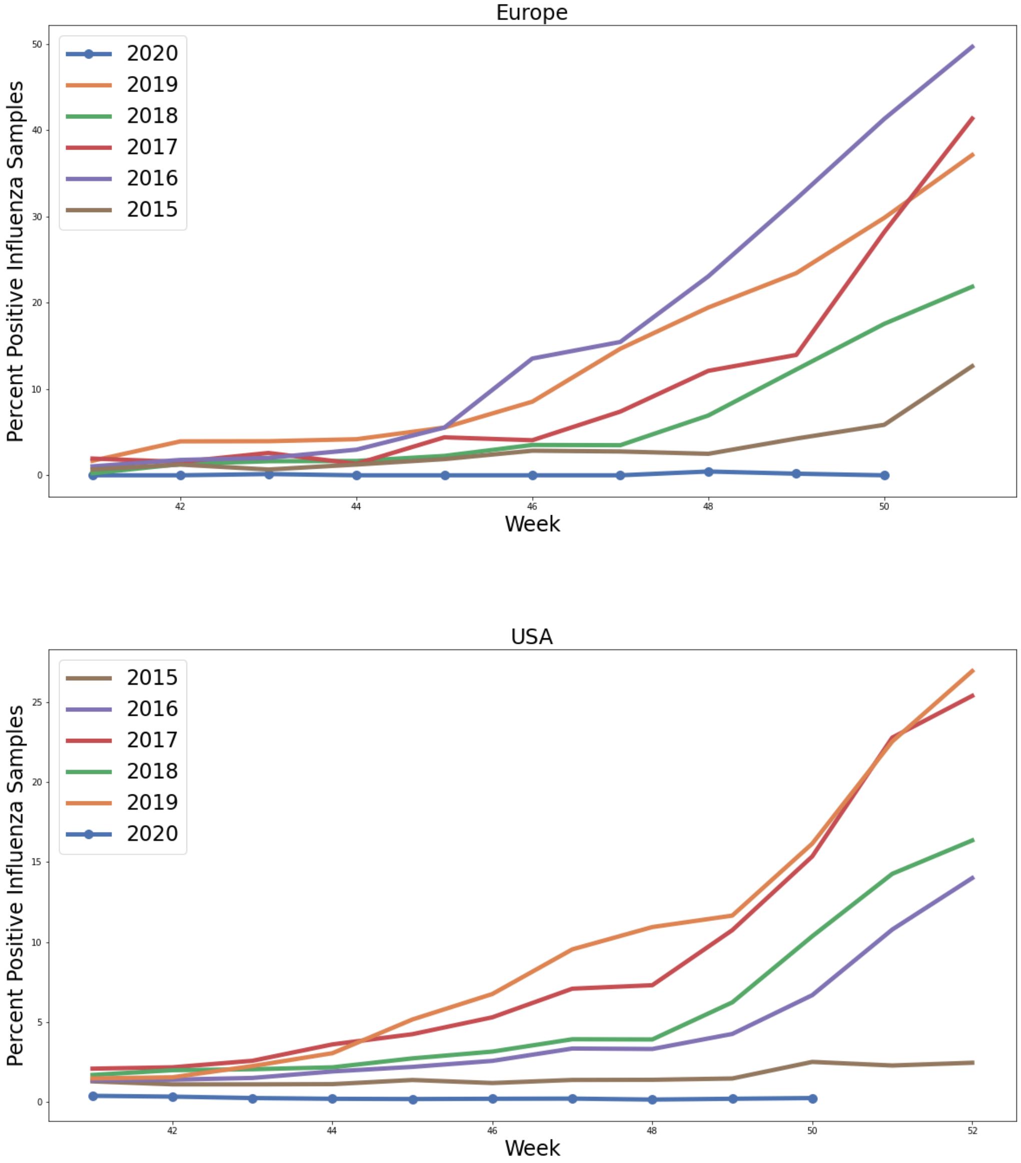


**Figure S4:** The proportion of respiratory samples that have tested positive for influenza remains low in the WHO Europe region and in the US.
